# A Mathematical Model for Stability Analysis of Covid like Epidemic/Endemic/Pandemic

**DOI:** 10.1101/2021.11.08.21265055

**Authors:** A.K. Awasthi, Sanjeev Kumar, Arun Kumar Garov

## Abstract

The transmission and spread of infectious disease like Covid-19 occurs through horizontal and vertical mode. The causative pathogens for such kind of disease may be bacterium, protozoa, virus or toxin. The infectious diseases like AIDS, SARS, MARS, Polio Plague, Bubonic Plague and Covid-19 have destroyed the social and economic structure of world population. The world scientific community adopts different mechanisms to model and analyse the population dynamics of infectious disease outbreaks. Mathematical Modelling is the most effective tool to take the informed decision about the containment, control and eradication of the pandemic. The main focus of Government and public health authorities is to design the strategy in destabilising the spread and impact of the infections. A series of models-SIR, SEIR, SEIRD, SEAIHCRD, SAUQAR has been under study to combat the Covid-19 since its inception. An effort has been made to design the model based on reproduction number, endemic equilibrium and disease-free equilibrium to curtail the impact of Covid-19 through stability analysis methods-Hurwitz stability criteria, Lyapunov Method and Linear Stability Analysis.

## 1. Introduction

History is replete with the epidemics/pandemic which have had long lasting effects on the human society. For example, the Black Death known as bubonic plague caused the death of as much as one third population of Europe. In the series of epidemics, Covid-19 has incurred great loss to human well-being and destroyed the social, economic structure since its inception. Whole of the world is experiencing the recursions of chaos created due to this viral disease. Cough, pneumonia, dyspnoea, exhaustion, fever, diarrhoea, inflection in lungs, respiratory problems are unexplained causes of Covid-19. A single infected person is transmitting the infection hundreds and thousands of the population. Closure of schools, colleges, restriction on interstate or international travelling, corona curfew, lockdown, reduction in social gathering are few of the outcomes of Covid. World health scientific community have developed vaccine for the protection of the masses. But it is also seen that Covid-19 Virus has been changing its form and creating fear among the society. Various studies have been proposed to expedite the project for containment and eradication of this disease. Mathematical models are evolving from time to time to assess the spread of disease and frame the policy to intervene this spread. Anwar Zeb. Et.al [1] considered the isolation of infected persons to reduce the risk of spread of covid-19 In this work-related stability of reproductive stability is discussed and found that if control the contact rate then the containment of Covid is possible. Pakwan Riyapan et.al. [2] analysed the transmission dynamics of Covid-19 with a case study in Bangkok Thailand. It is proved that disease free equilibrium is globally asymptotically stable if basic reproduction number (R_cvd_) is less than one and endemic equilibrium occurs if R_cvd_ >1. Idris Ahmed et.al. [3] used ODE and fractional differential equation to describe the outbreak of Covid-19. In this model disease equilibrium point (E_0_) is found to be locally asymptotically stable, whenever the basic reproduction number R_0_<1 and endemic equilibrium (E_1_) is obtained as globally asymptotically stable whenever R_0_>1. Faical Nairo et.al. proposed compartmental mathematical model with the transmissibility of super-spreader individual on Covid-19 and studied the local stability of the disease-free equilibrium in terms of basic reproduction number with a case study in Wuhan. Mohammed A. Abaoud et.at. [5] applied the Caputo/fractional derivatives to understand and give more insight about the transmission dynamics of coronavirus with numerical simulation. Vipin Tiwari et.al. studied five compartmental model SEIRD and predicted the Covid-19 dynamics peak value under the impact of lockdown in India. [7] Avaneesh Singh et.al. extended SEIR model to SEAIHCRD which includes asymptomatic infected, hospitalised, critical people with dead compartment. In this model author computed the infection rate, recovery rate, case fatality rate by taken into account the various parameters like age group, hospital beds, proper social distancing etc. Masaki et.al.[8] constructed SIIR (Susceptible, Infection, Incubation, Recovered) Model and described the spread of infection with the consideration of novel Covid-19. It is found that herd community is more susceptible to disease in SIIR as that of in SIR model. The main point in this study is that after infection disease carriers can spread during incubation period, which is very difficult to handle. Constanttinose I, Siettos et.al. [9] categorised epidemiological model into three parts-Statistical, Mathematical mechanistic state space and machine learning based. This study is based on chronological order epidemics from Cholera in 1854 at London to global AIDS epidemics. A better understanding of the signature features of epidemic outbreaks from real outbreak data and different mathematical modelling approach could lead to substantial improvement in our ability to forecast the epidemics [10]. Epidemic growth profiles range from sub exponential to exponential growth across the number of epidemic outbreaks such influenza, smallpox, measles, HIV/AIDS and Ebola. Different mathematical techniques have been applied to characterise the epidemics growth dynamics. Successful efforts in improving disease transmission modelling toward improved disease forecasting will also have an impact on refining preparedness and contingency interventions plans to confront infection disease threats.

## 2. Mathematical formulation of Epidemiological Model SVEIR

The whole population is divided mainly into five distinct epidemiological subclasses of individual- (S)susceptible population, (V)Vaccinated Population, (E)exposed population, (I)infected population, and (R)recovered population. It is observed in common practice that when the susceptible population is vaccinated, some of the vaccinated population is not getting the benefit of vaccination and some part of the population is recovering directly without any kind of infection. The chances of vaccinated people getting exposed always lies there. Every new born is equally likely to get infected therefore both horizontal and vertical transmission is considered in this work. The total population size at time t is denoted by N(t), with

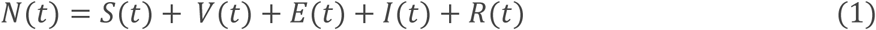

Let *bN* is total number of new born with natural birth rate b, *pbI* is the number of new born who are infected at birth, *bN* − *pbI* is the number of healthy but susceptible new born.

The following SVEIR epidemic model along with transfer diagram for migration/immigration and removal is shown in Figure-1. And set of ordinary differential equations are established to analyse the stability of disease-free equilibrium and endemic equilibrium.

**Figure-1:**
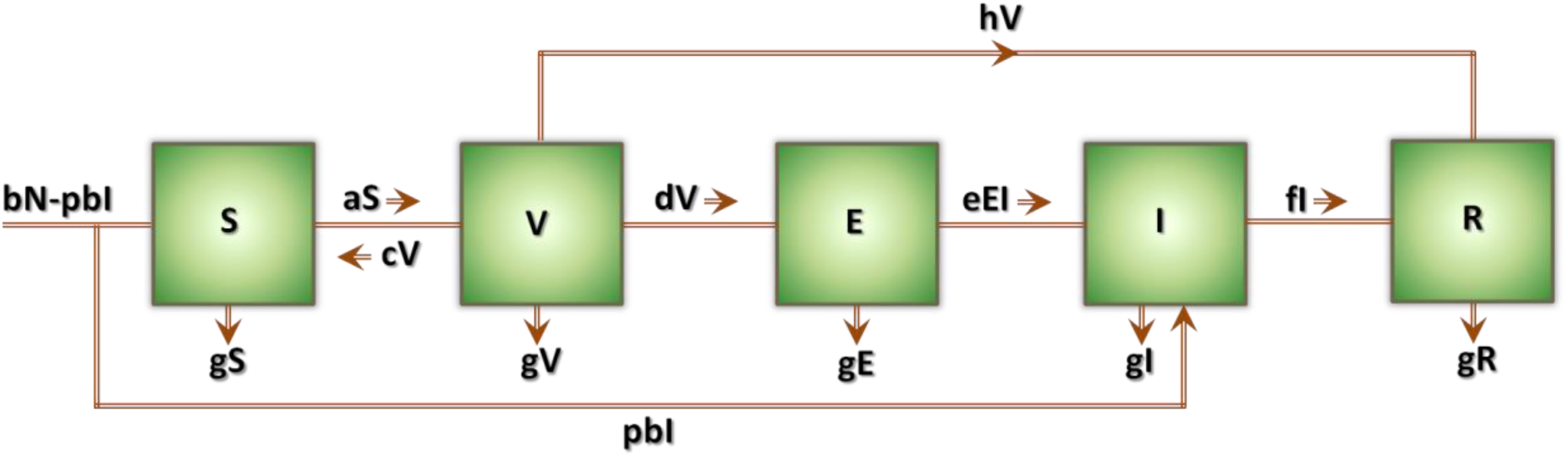
Transfer diagram for SVIER model with migration/immigration and removal

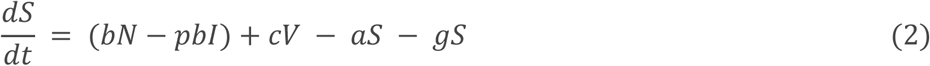

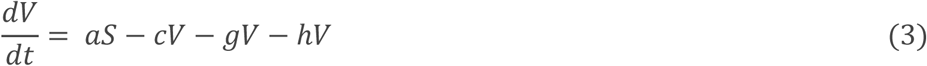

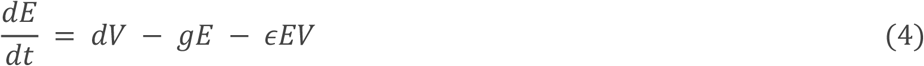

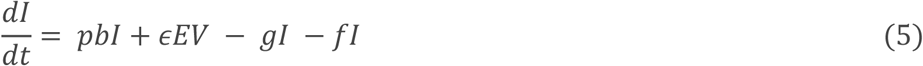

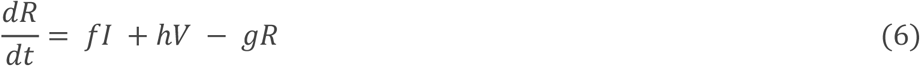

where b is natural birth rate, *a* is the rate of vaccination for susceptible population, and c is the rate at which vaccinated population again enters into susceptible population, p is the fraction of born infected, g is the natural death rate, d is rate of vaccination, *ϵ* is rate of infection to infected class, *f* is the rate of recovery and *h* is the rate at which vaccinated class is directly recovered without getting infected.

## 3. Next Generation Matrix and Reproduction Number/ Equivalent threshold parameter

In this section, we determine the basic reproduction number R_0_ and obtain the existence of the disease-free equilibrium (DFE) and the endemic equilibrium (EE) of system (2-6). Summing up the five equations of system (2-6) we get

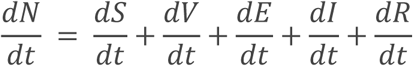

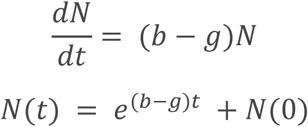

Therefore, from biological considerations, we study system (2-6) in the following feasible region

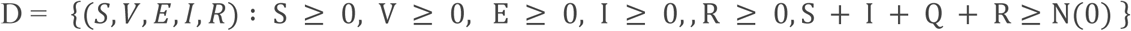

In a literature review, it is found that authors have derived the equivalent threshold parameter also knowns as reproduction number or reproduction ratio when more than one class of infectives are involved. Diekmann et al. (1990), introduced the next generation method to derive the Reproduction number (R_0_), where the population has been divided into discrete and disjoint classes. In the next generation method, R_0_ is defined as the spectral radius of the next generation operator. The formation of the operator involves determining two compartments, infected and non-infected, from the model. In this section, we outline the steps needed to find the next generation operator in matrix notation (assuming only finitely many types), and then employ this method for a susceptible–vaccinated-exposed– infectious–recovered (SVEIR) model. Consider for a set of n compartments, out of which m are infected. Let us define the vector 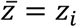 where *z*_*i*_ denotes the number or proportion of individuals in the ith compartment. Let 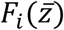 be the rate of appearance of new infections in the i^th^ compartment. And let 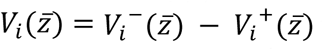 where *V*_*i*_^−^ is the rate of transfer of individuals into compartment i by all other means and *V*_*i*_^+^ is the rate of transfer of individuals out of the i th compartment. The difference 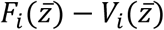 gives the rate of change of *z*_*i*_. Note that F_i_ should include only infections that are newly arising, but does not include terms which describe the transfer of infectious individuals from one infected compartment to another. Assuming that F_i_ and V_i_ meet the conditions outlined by Diekmann et al. (1990) and van den Driessche & Watmough (2002), we can form the next generation matrix (operator) FV^-1^ from matrices of partial derivatives of F_i_ and V_i_. Specifically, 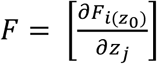 and 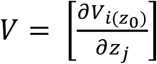; where i, j =1,2,3…, m and where *z*_0_ is the disease-free equilibrium. The entries of FV^-1^ give the rate at which infected individuals in *z*_*j*_ produce new infections in *z*_*i*_, times the average length of time an individual spends in a single visit to compartment j. R_0_ is given by the spectral radius (dominant eigenvalue) of the matrix FV^-1^.

The model dynamic defined by the equations (1-6) for the SVIER model gives us *F* and *V* as follows.

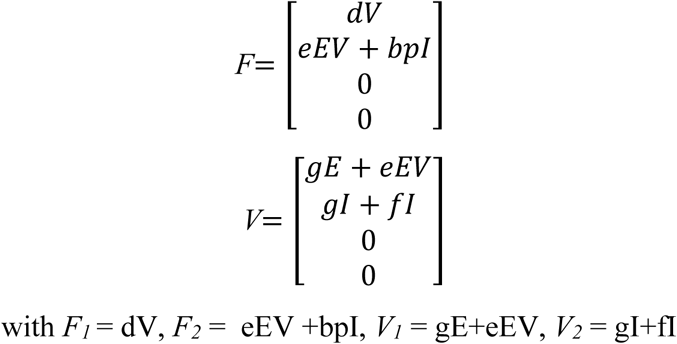

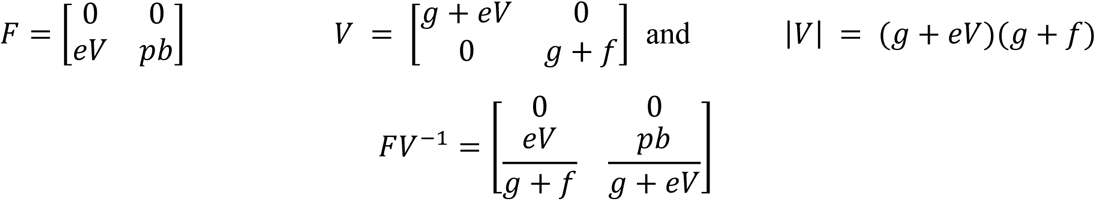

Now, the spectral radius of FV^-1^ is the reproduction number,

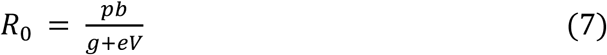

## 4. Disease Free and Endemic Equilibrium

In this section we will obtain the disease free and endemic equilibrium point for the system described by (2-6).

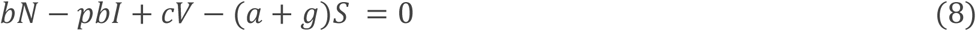

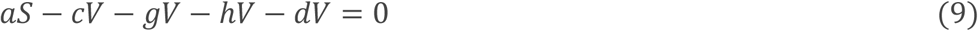

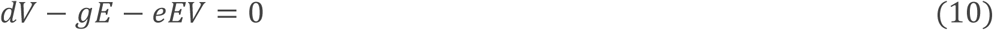

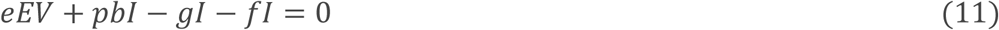

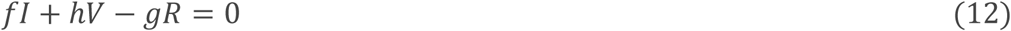

On solving equations (8-12), we get the disease-free equilibrium (DFE) point Z_0_ (0,0,0,0,0) and the Jacobian matrix for the above system is given by: -

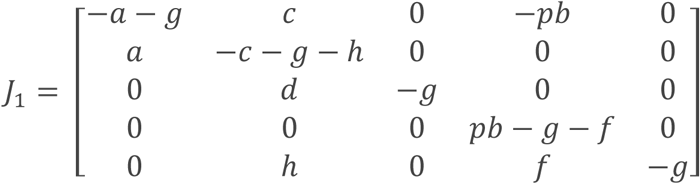

The eigen values for the above matrix are: -

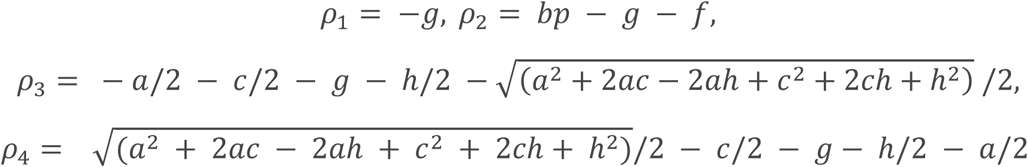

For all the given parameters *ρ*_*i*_ < 0 *for i* = 1,2,3,4. Therefore, disease free equilibrium is asymptotically stable.

The endemic equilibrium (EE) point z_i_ for the system (8-12) is found to be: -

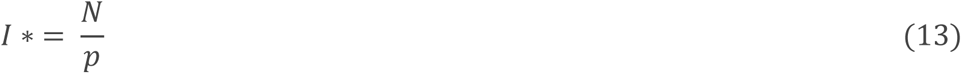

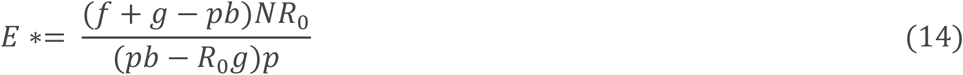

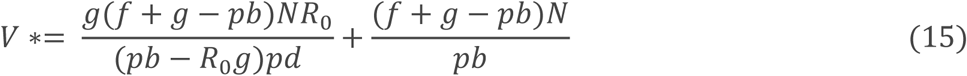

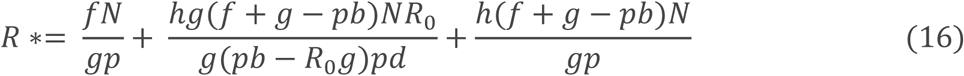

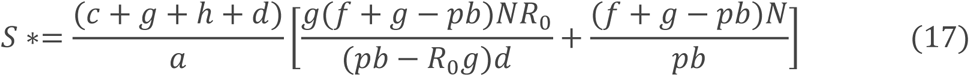

The Jacobian matrix for the system (2-6) is given by: -

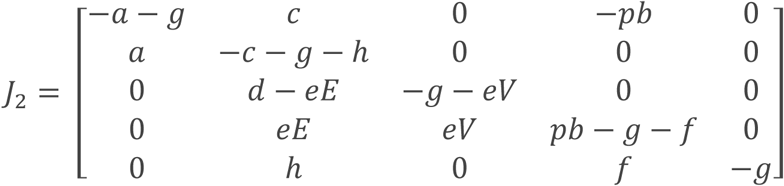

It is cumbersome to obtain the analytical expression for the eigen values of J_2_ with the use (13-17). Therefore, we have made an attempt to discuss the result with numerical analysis.

## 5. Result and Discussion

In the study of SVEIR model it is found that disease free equilibrium (DFE) is asymptotically stable for all values of the parameters a, b, c, d, e, f, g, h. To analyse the stability of endemic equilibrium we have taken the following examples: -

### Example 1 (DFE): -

Let a=0.7; b=0.3; c=0.2; d=0.35; e=0.45; h=0.8; f=0.5; V=75; N=100; p=0.4; E=50; g=0.3;

**Reproduction Number**

R_0_ = 0.0035 < 1

### Theorem

If R_0_ < 1, the disease-free equilibrium of system is locally asymptotically stable. If R_0_ > 1, the disease-free equilibrium is unstable.

In our example, R_0_ = 0.0035 < 1 therefore, the system should be stable.

Now the eigen values corresponding to the Jacobian matrix are found to be: -’

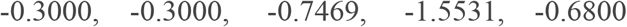

which are less than zero. Hence the given system is stable.

### Example 2 (EE): -

a=0.99; b=5; c=0; d=0.2; e=0.92; h=0.7; f=0.97; V=1; N=100; p=0.999; E=90; g=0.998;

### Theorem.

If R0 > 1, the endemic equilibrium of system is globally asymptotically stable.

**Reproduction Number**

R_0_ = 1.4705 > 1

And the eigen values corresponding to the Jacobian matrix are: -

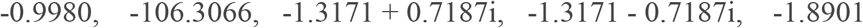

All of the eigen values are either negative or have negative real part, hence the given system is stable that is disease is not going to occur for large duration and it will end up initial hazards.

So, the health policy makers can keep the check over the parametric values and take the valuable decision to curb the invasion and outbreak of pandemic like COVID-19.

### 5.1 Stability Analysis based on Routh Hurwitz criteria

It is very much possible that Jacobian matrix of system (2-6) remain inconclusive for the stability analysis, in that case we can use Routh Hurwitz criteria in following way: -

Let the characteristic equation of Jacobian matrix is defined by: -

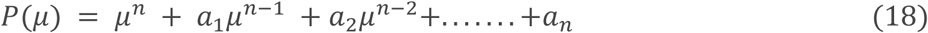

Consider *M*_1_ = *a*_1_,

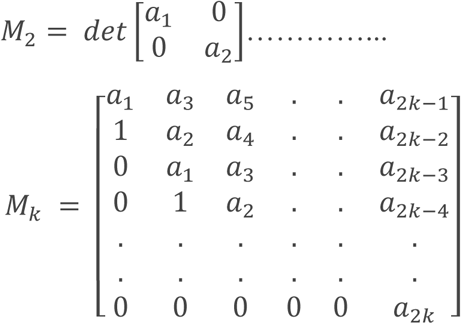

Where *a*_*i*_ = 0 if *i* > *n*. Then the roots of *P*(*μ*) have negative real parts if and only if *M*_*k*_ > 0 for all *k* = 1,2,3……*n*. For example, let n = 2

Then we have

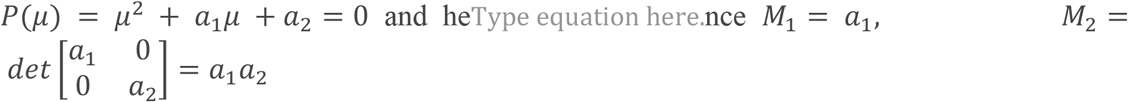

Thus, for n = 2 the necessary and sufficient condition for having roots with negative parts are *a*_1_ > 0, *a*_2_ > 0. Similarly for higher order matrices we can have different conditions on *a*_*i*_, to have negative real parts of the roots of characteristics equation, based upon which stability of the system can be decided.

### 5.2 Stability Analysis based on Lyapunov function

Basically, Lyapunov’s Direct Method is used to describe the stability of linear, non-linear mechanical, electrical or physical system. In this description, the total energy of the system is dissipated and eventually the system the system is reduced to its lowest level known equilibrium point.

Definition: If in the given domain, the function F(x) is positive definite and has continuous partial derivatives, and if its time derivative along any state trajectory of system is negative semi definite that is 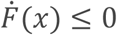, Then F(x) is said to be Lyapunov function. And the point for which this function exist is said to be stable. The stability is Asymptotic Global Stable if 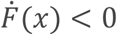.

In this section we establish the global stability with the help of Lyapunov function for the SVEIR model. Consider the endemic equilibrium point Z*(S*, V*, E*, I*, R*) of the system (2-6). For the above system we construct the following positive definite function.

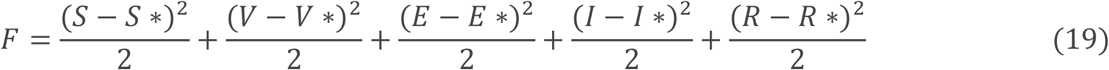

Clearly, *F*: *R*_+_^5^ → *R* is continuously differentiable function. It is easy to see that F*(S*, V*, E*, I*, R*) = 0 and F*(S*, V*, E*, I*, R*) > 0 for all (S*, V*, E*, I*, R*) ≠ (S, V, E, I, R). Now differentiating (19) w.r.t ‘t’ we get the following equation.

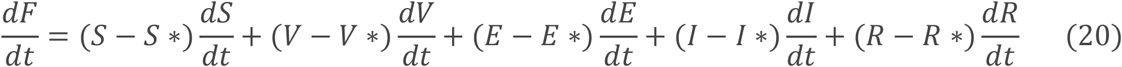

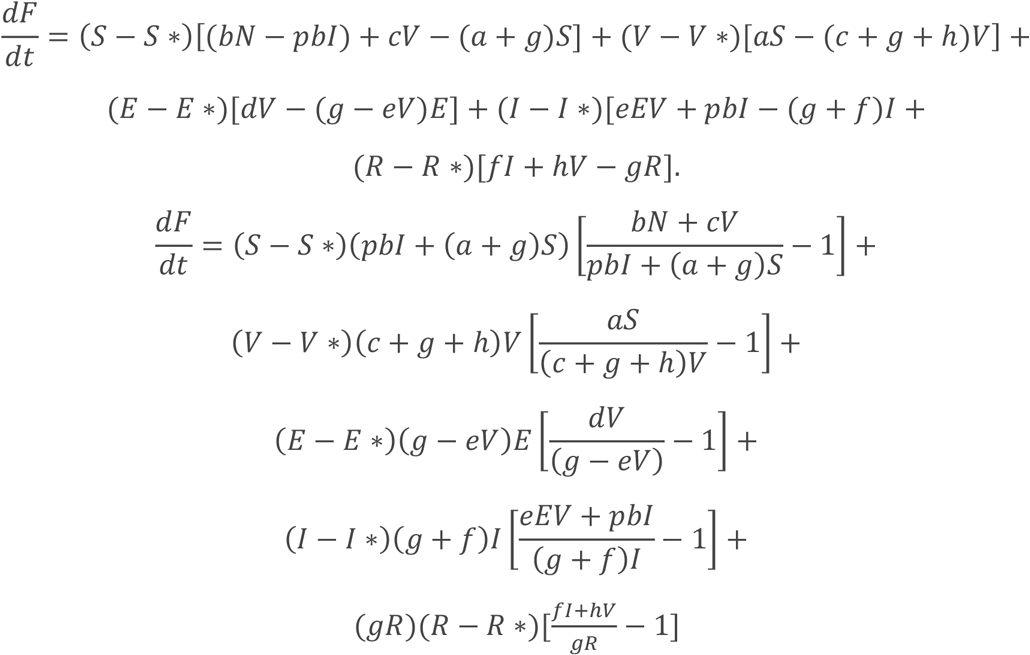

Now 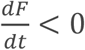, at Z* if the following conditions hold true.

a. 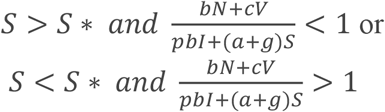
b. 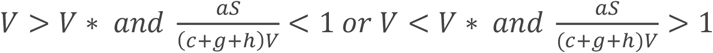
c. 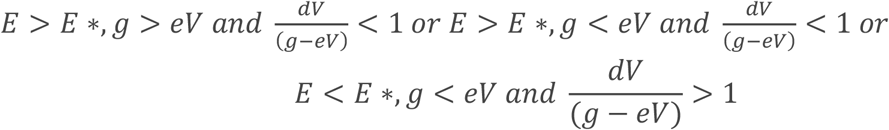
d. 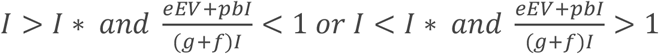
e. 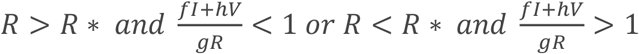

Thus, the SVEIR model (2-6) is globally asymptotically stable with R_o_ > 1 along with the conditions (a-e).

## 6. Conclusion

In this paper, we have formulated an SVEIR epidemic model with vaccination, elimination, and exposed and infectious population classes, and studied the dynamics of this disease model by means of both theoretical and numerical ways. For this model, we defined the basic reproduction number R_0_ which completely determines the dynamical behaviour of system (2-6). When R_0_ < 1, as is shown in Theorem 1, the disease-free equilibrium is globally asymptotically stable and the disease always dies out eventually. When R_0_ > 1, Theorem 1 tell us that the unique endemic equilibrium is globally asymptotically stable and the disease persists at the endemic equilibrium level if it is initially present. Some numerical examples are taken to illustrate the analysis results. Finally, we discussed and analysed the characteristics of different control strategies according to the basic reproductive number R_0_.

## Data Availability

All data produced in the present study are available upon reasonable request to the authors
All data produced in the present work are contained in the manuscript

http://www.google.com

http://www.google.com

## Notes

### Competing Interest Statement

The authors have declared no competing interest.

### Clinical Protocols

http://www.google.com

http://www.google.com

http://www.google.com

### Funding Statement

This study did not receive any funding

